# Performance of open-source large language models on nephrology self-assessment program

**DOI:** 10.64898/2026.04.16.26348910

**Authors:** Meysam Ahangaran, Shuyue Jia, Shlok Chitalia, Ambarish Athavale, Jean M. Francis, Michael William O’Donnell, Santhoshi Rupa Bavi, Uma Datta Gupta, Vijaya B. Kolachalama

**Affiliations:** Department of Medicine, Boston University Chobanian & Avedisian School of Medicine, Boston, MA, USA; Department of Electrical & Computer Engineering, Boston University, Boston, MA, USA; Division of Nephrology-Hypertension, University of San Diego, California, San Diego, California, USA; Department of Computer Science and Faculty of Computing & Data Sciences, Boston University, MA, USA, Wentworth Institute of Technology, School of Engineering, Boston, MA, USA; Boston Medical Center, Section of Nephrology, Boston, MA, USA

## Abstract

**Background:** Large Language Models (LLMs) have demonstrated strong performance in medical question-answering tasks, highlighting their potential for clinical decision support and medical education. However, their effectiveness in subspecialty areas such as nephrology remains underexplored. In this study, we assess the performance of open-source LLMs in answering multiple-choice questions from the Nephrology Self-Assessment Program (NephSAP) to better understand their capabilities and limitations within this specialized clinical domain.

**Methods:** We evaluated the performance of five open-source large language models (LLMs): PodGPT which a podcast-pretrained model focused on STEMM disciplines, Llama 3.2-11B, Mistral-7B-Instruct-v0.2, Falcon3-10B-Instruct, and Gemma-2-9B-it. Each model was tested on its ability to answer multiple-choice questions derived from the NephSAP. Model performance was quantified using accuracy, defined as the proportion of correctly answered questions. In addition, the quality of the models explanatory responses was assessed using several natural language processing (NLP) metrics: Bilingual Evaluation Understudy (BLEU), Word Error Rate (WER), cosine similarity, and Flesch-Kincaid Grade Level (FKGL). For qualitative analysis, three board-certified nephrologists reviewed 40 randomly selected model responses to identify factual and clinical reasoning errors, with performance summarized as average error ratios based on the proportion of error-associated words per response.

**Results:** Among the evaluated models, PodGPT achieved the highest accuracy (64.77%), whereas Llama showed the lowest performance with an accuracy of 45.08%. Qualitative analysis showed that PodGPT had the lowest factual error rate (0.017), while Llama and Falcon achieved the lowest reasoning error rates (0.038).

**Conclusions:** This study highlights the importance of STEMM-based training to enhance the reasoning capabilities and reliability of LLMs in clinical contexts, supporting the development of more effective AI-driven decision-support tools in nephrology and other medical specialties.

## Introduction

Large language models (LLMs) are artificial intelligence (AI) systems designed to process, understand, and generate human-like text. Leveraging extensive training on vast datasets, models such as OpenAIs GPT-4, along with other proprietary and open-source alternatives, have exhibited remarkable performance across various domains. In medicine, LLMs have been increasingly evaluated for their ability to answer multiple-choice questions, showcasing their potential for knowledge-based assessments. As their applications continue to expand, LLMs are emerging as transformative tools in natural language processing, with significant implications for research, education, and problem-solving. In this study, we assessed the performance of PodGPT^1^, a recently introduced LLM in answering nephrology multiple-choice test questions and compared its accuracy against four open-source LLMs including Llama 3.2-11B (September 2024)^2^, Mistral-7B-Instruct-v0.2 (December 2023), Falcon3-10B-Instruct (December 2024)^3^, and Gemma-2-9B-it (December 2024)^4^.

PodGPT was developed using a diverse corpus of text extracted from publicly available podcast data from science, technology, engineering, mathematics and medical (STEMM) disciplines. It also leverages retrieval-augmented generation (RAG) to incorporate the latest medical evidence from high-impact literature, including *The New England Journal of Medicine* (NEJM). As NephSAP usually gives nephrologists a summary of the most recent literature and publications related to a topic published within two years prior to the release of the review, the superior performance of PodGPT may suggest its enhanced access to up-to-date medical content, potentially contributing to its stronger results compared to other systems. Additionally, we assessed model reasoning by comparing generated justifications to the reference standard using NLP-based similarity metrics (**Figure 1**).

**Figure 1.**
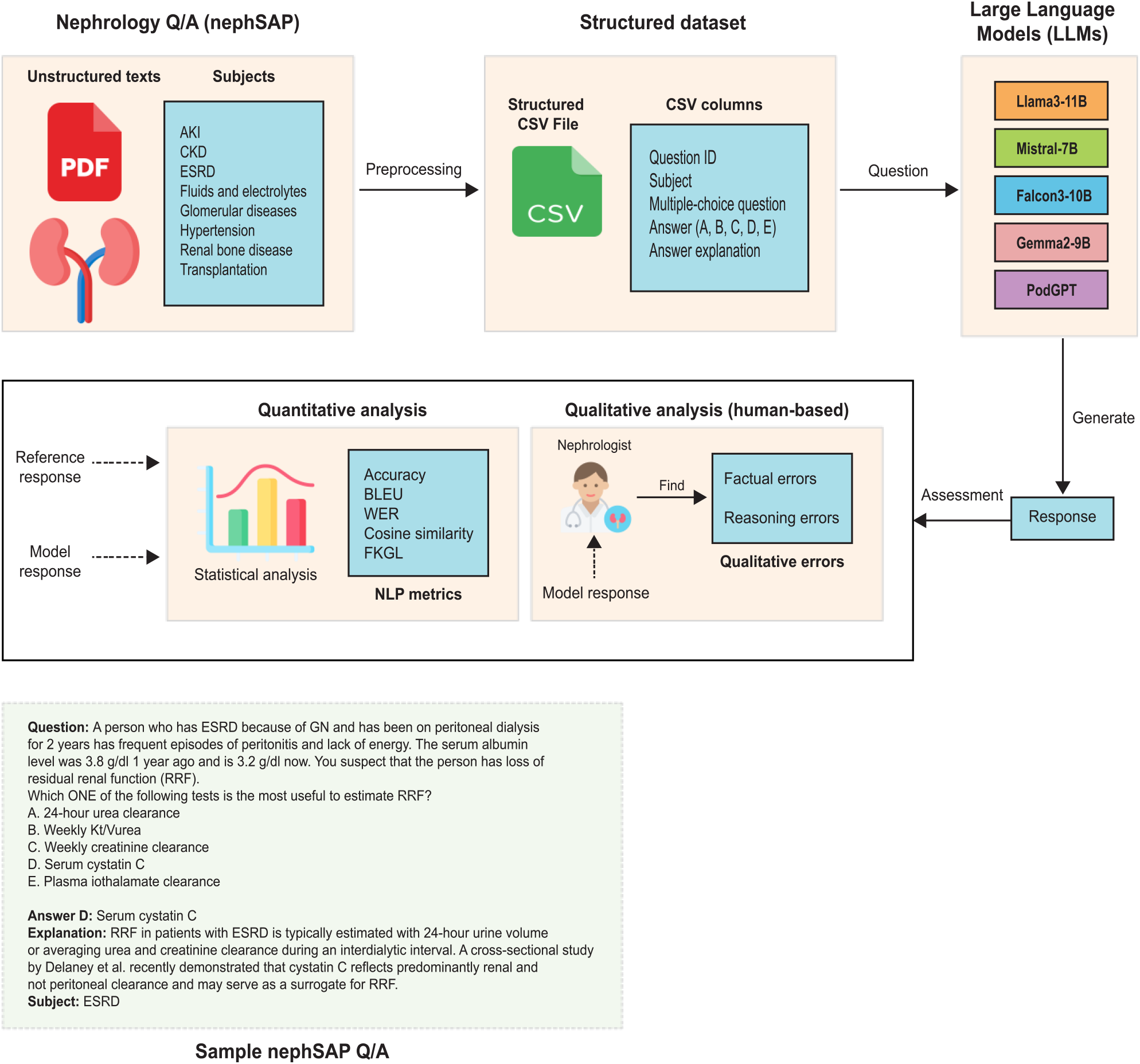
Study overview. The schematic presents the overall study workflow, encompassing data collection, preprocessing, and utilizing Large Language Models (LLMs) for nephrology Self-Assessment Program (nephSAP) multiple-choice questions.

## Methods

### Dataset

The dataset utilized in this study comprises 528 multiple-choice questions from the nephrology self-assessment program (nephSAP), each accompanied by explanations and reference citations. nephSAP is designed to help learners systematically evaluate their strengths and weaknesses across various nephrology domains^5^. Each question follows a structured format, presenting a clinical scenario followed by a prompt requiring the selection of the correct answer from multiple choices. The dataset covers eight key nephrology subjects, with the following questions distribution: acute kidney injury (AKI) and critical care nephrology (N=45), chronic kidney disease (CKD) and progression (N=75), end stage renal disease (ESRD) and dialysis (N=70), fluids, electrolytes, and acid-base (N=45), glomerular, vascular, and tubulointerstitial disease (N=104), hypertension (N=75), renal bone disease, disorders of divalent ions, and nephrolithiasis (N=40), and transplantation (N=74). To prepare the dataset for evaluation, we extracted a structured CSV file from raw text using a parsing script. The final CSV file comprised of five columns: identifier, subject, question, answer, and explanation. This structured dataset was then used to assess the performance of LLMs on nephSAP benchmarks.

### Evaluation of language models

To assess the quality of the answers generated by the LLMs based on NLP metrics, we developed an automated script to facilitate the analysis of model outputs. For each model, we calculated the percentage of correctly answered questions. Additionally, we employed the BiLingual Evaluation Understudy (BLEU) metric, implemented via the NLTK library^6^, to compare the quality of generated answers with the reference standard. The BLEU score ranges from 0 to 1, with 0 indicating poor quality and 1 reflecting perfect similarity to the reference standard. We also used word error rate (WER)^7^ and cosine similarity^8^ to compare between the models output and the reference explanation. WER ranges from 0% to 100%, with lower values indicating higher accuracy, while cosine similarity ranges from 0 to 1, with a higher value indicating greater similarity to the reference standard. In addition, we calculated the Flesch-Kincaid Grade Level (FKGL) to quantify the readability of model-generated responses. FKGL estimates the U.S. school grade level required to comprehend a given passage, with lower scores indicating simpler, more accessible language. Finally, we evaluated the models ability to answer difficult questions (161 out of 528), defined as those where only one or fewer models provided correct answers.

To evaluate response quality through human assessment, three board-certified nephrologists independently reviewed responses generated by five models to identify *factual* and *reasoning* errors. Factual errors were defined as inaccurate medical information in the model-generated responses, whereas reasoning errors referred to flawed clinical reasoning, including incorrect diagnostic interpretation or inappropriate treatment recommendations. The nephrologists were asked to review 40 random responses across eight nephrology subject areas and identify sentences containing factual or reasoning errors.

## Results

### Quantitative analysis

Among the evaluated models, PodGPT achieved the highest overall accuracy (0.65) by correctly answering 342 out of 528 questions. In contrast, Llama recorded the lowest accuracy (0.45), followed by Mistral (0.50), Gemma (0.52), and Falcon (0.57). When performance was stratified by question difficulty, Gemma showed the lowest accuracy for difficult questions (0.06), while PodGPT again led with 0.20 accuracy (**Figure 2A**).

**Figure 2.**
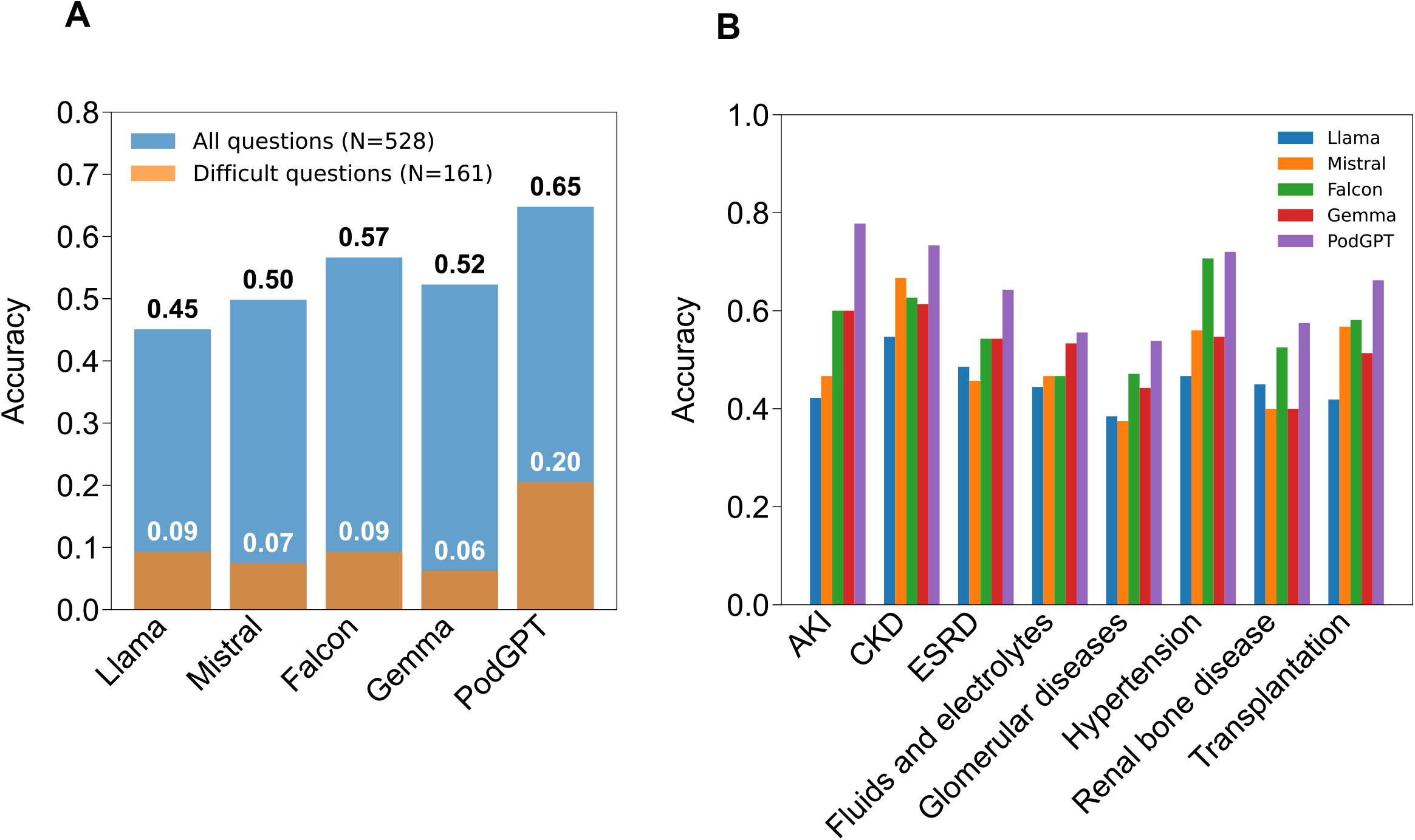
Comparison of large language model (LLM) performance on overall and subject-specific accuracy. **(A)** Accuracy of five LLMs (Llama, Mistral, Falcon, Gemma, and PodGPT) was evaluated on 528 total questions and a subset of 161 “difficult” questions, defined as those correctly answered by at most one model. (B) Subject-specific accuracy was evaluated across eight core nephrology domains: acute kidney injury (AKI), chronic kidney disease (CKD), end-stage renal disease (ESRD), fluids and electrolytes, glomerular diseases, hypertension, renal bone disease, and transplantation.

Subject-specific performance revealed that PodGPT consistently outperformed other models across all eight nephrology domains (**Figure 2B**). Llama had the lowest accuracy in five of the subjects (AKI, CKD, fluids and electrolytes, hypertension, and transplantation), while Mistral performed the worst in ESRD, glomerular diseases, and renal bone disease. Among all subject areas, glomerular diseases were the most challenging, with the lowest average model accuracy (0.44), while CKD had the highest overall accuracy (0.64).

### Assessing the quality of model explanations with NLP metrics

The quality of answers generated by the LLMs was evaluated using BLEU, WER, cosine similarity, and FKGL metrics, with 95% confidence intervals (**Figure 3**). BLEU scores ranged from 0.032 for Llama to 0.074 for PodGPT, indicating notable differences in language generation quality across models. WER scores were lowest for Mistral (1.17%) and highest for Llama (3.64%), suggesting differences in transcription or speech alignment performance. When evaluating semantic quality using cosine similarity, scores were moderate across the board, with Falcon achieving the highest similarity score (0.666) and Llama the lowest (0.583). Analysis of the FKG score revealed that Llama produced the least complex explanations, with a score of 12.7, while the ground truth exhibited the highest complexity, with a score of 16.23. The FKG scores of the remaining models fell within the range of the lowest and highest values, with slight variations (**Table 1**).

**Table 1.**
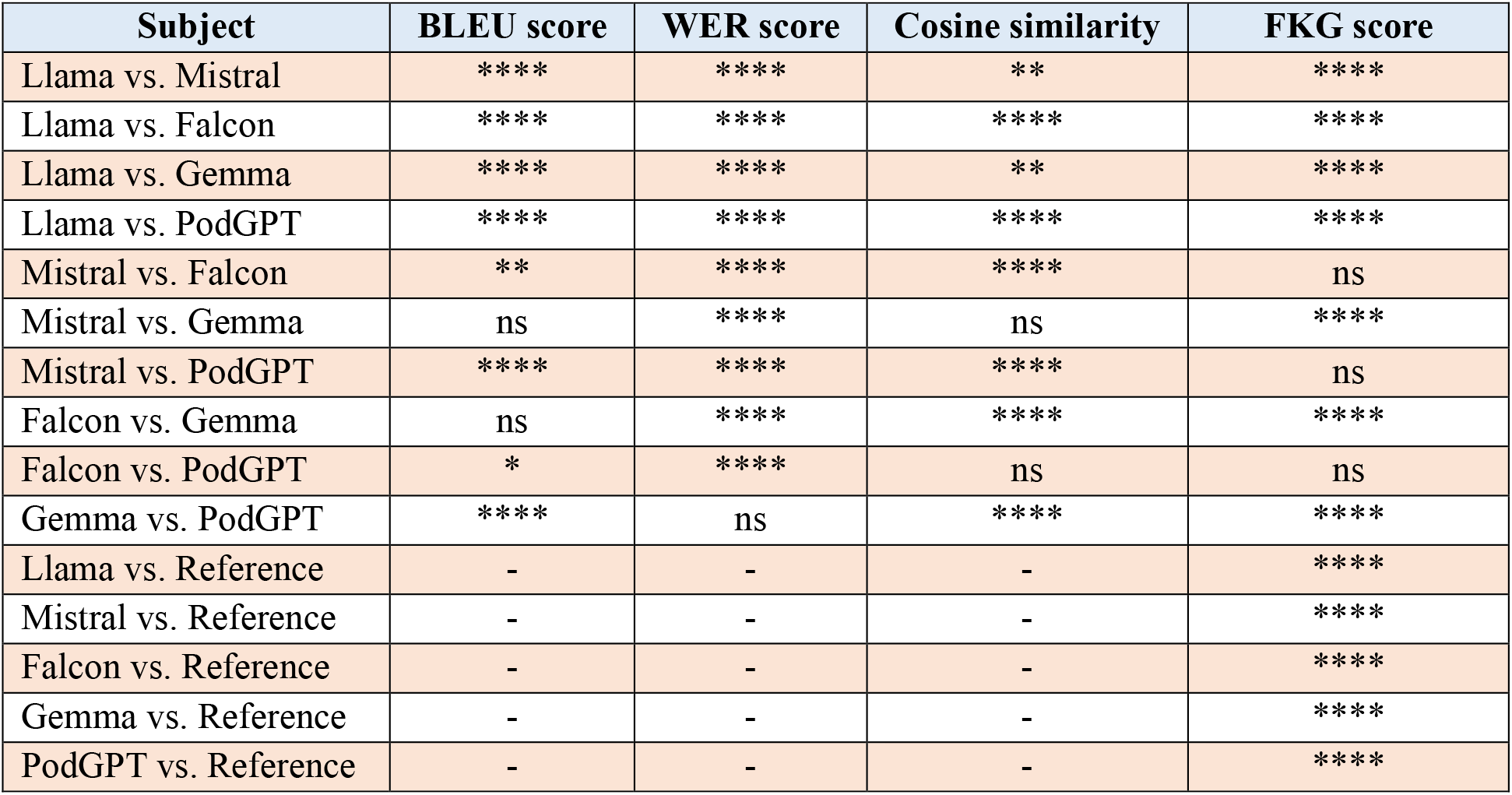
Statistical comparisons between large language models. P-values were derived using the Kruskal-Wallis test, followed by Bonferroni-adjusted pairwise comparisons. Significance levels are denoted as *ns* (not significant) for P > 0.05, *P ≤ 0.05, **P ≤ 0.01, ***P ≤ 0.001, ****P ≤ 0.0001 (see **Figure 3**).

**Figure 3.**
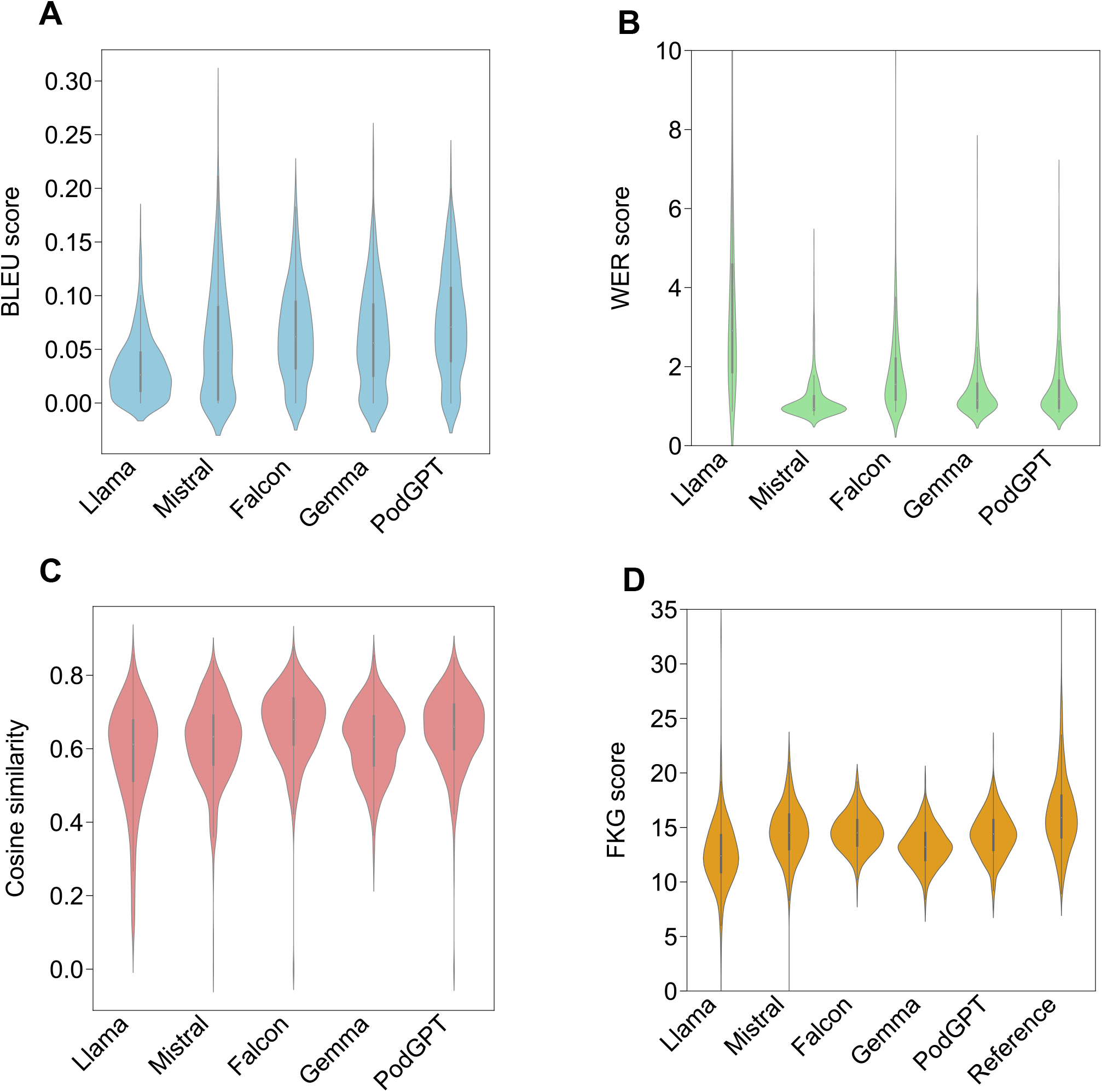
Evaluation of language models across accuracy, BLEU score, word error rate, cosine similarity, and readability. (A) BLEU scores reflect the degree of n-gram overlap between model-generated answers and the reference answers. (B) Word error rate (WER) quantifies discrepancies at the word level between generated and reference answers. (C) Cosine similarity scores were computed between sentence embeddings of generated and reference answers. (D) Flesch-Kincaid Grade Level (FKGL) estimates the readability of model responses. Error bars represent 95% confidence intervals.

### Qualitative analysis

To evaluate response quality via expert assessment, three board-certified nephrologists independently examined responses produced by five models for factual inaccuracies and errors in clinical reasoning. For this analysis, five questions were randomly selected from each of the eight nephrology subject areas in NephSAP, yielding a total of 40 responses. Nephrologists reviewed each response to identify sentences containing factual or clinical reasoning errors. To quantify error ratio, we calculated the proportion of erroneous content by dividing the total number of words associated with each error type by the total number of words in the corresponding response, thereby obtaining average factual and reasoning error ratio for each model. As shown in **Figure 4**, PodGPT demonstrated the lowest factual error rate (0.017), whereas Llama and Falcon exhibited the lowest reasoning error rates (0.038). In contrast, Mistral showed the poorest performance in both categories, with factual and reasoning error rates of 0.089 and 0.079, respectively.

**Figure 4.**
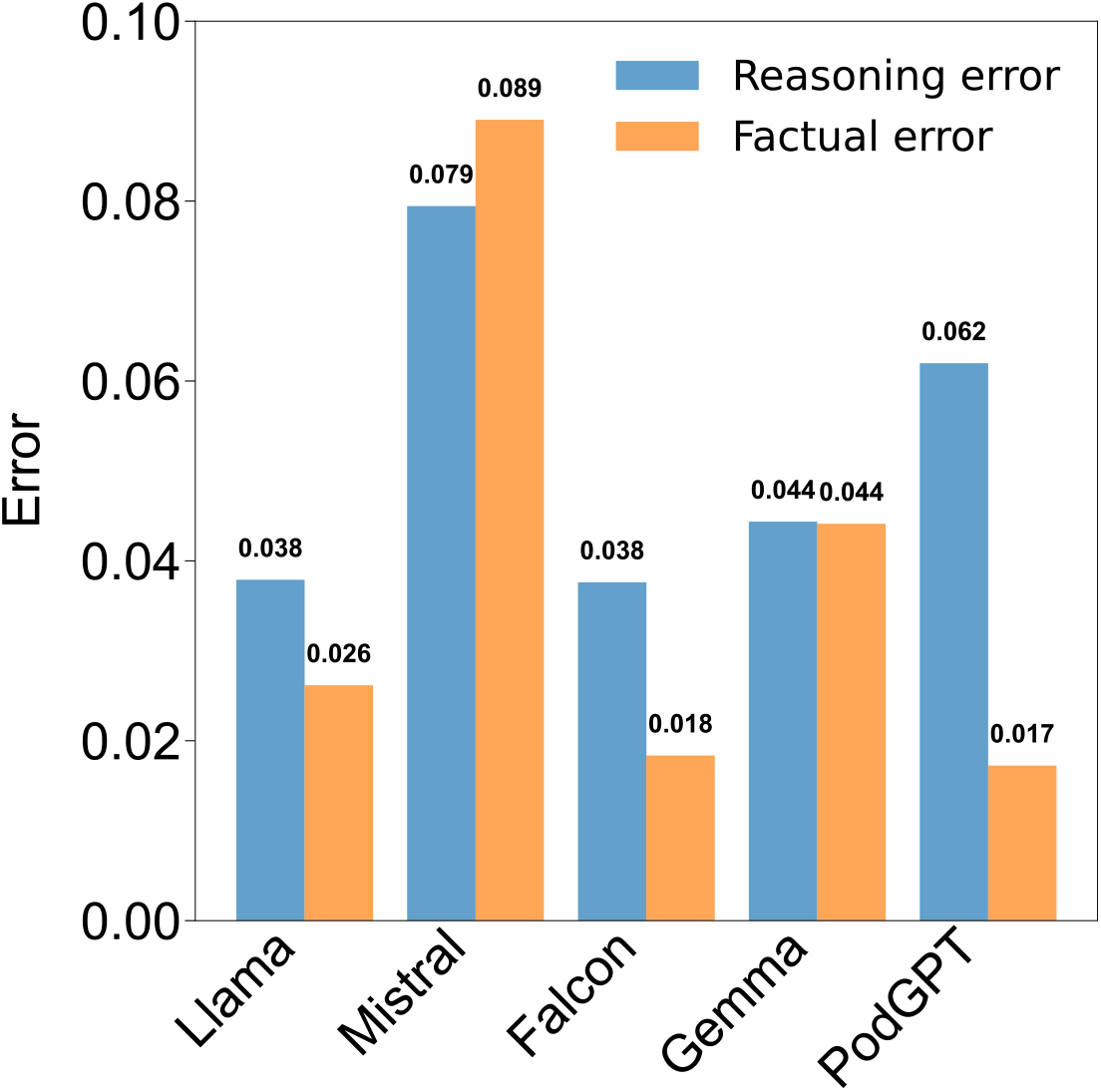
Human evaluation of model responses by nephrologists. Three nephrologists assessed 40 randomly selected responses (five questions from each of eight nephrology subject areas) to identify factual and reasoning errors. Bars represent the proportion of erroneous words relative to total response length for each model. PodGPT showed the lowest factual error rate (0.017), while Llama and Falcon demonstrated the lowest reasoning error rate (0.038). Mistral exhibited the highest factual and reasoning error rates (0.089 and 0.079, respectively).

## Discussion

Our results demonstrate that LLMs vary in their ability to answer nephrology multiple-choice questions, with PodGPT outperforming open-source alternatives across all nephrology subjects. However, performance gaps remain, particularly in subspecialty areas such as glomerular diseases, underscoring the need for further model refinement. As AI-driven education and decision support tools continue to evolve, healthcare systems may integrate LLMs into clinical workflows and physician training programs. Subspecialty fields such as nephrology stand to benefit from enhanced AI-driven knowledge dissemination and personalized medical training. Future work should focus on optimizing real-time updates to align with evolving medical guidelines. In addition, future research could explore the capabilities of commercially available LLMs such as ChatGPT and OpenEvidence for nephrology education, self-assessment and clinical support.

## Data Availability

The NephSAP question answer dataset analyzed in this study is maintained by the American
Society of Nephrology (ASN) and is accessible through the NephSAP program to authorized users and licensed nephrologists.

https://www.asn-online.org/education/nephsap

## Data availability

The NephSAP question–answer dataset analyzed in this study is maintained by the American Society of Nephrology (ASN) and is accessible through the NephSAP program (https://www.asn-online.org/education/nephsap) to authorized users and licensed nephrologists.

## Code availability

Python scripts for quantitative and qualitative analyses, including NLP metrics computation and figures generation, are available on GitHub at https://github.com/me-ahangaran/nephSAP-LLM.

## Funding

This project was supported by grants from the National Institute on Agings Artificial Intelligence and Technology Collaboratories (P30-AG073104, P30-AG073105), and the National Institutes of Health (R01-HL159620, R01-AG083735, R01-AG062109, and R01-NS142076).

## Conflict of interest

V.B.K. is a co-founder and equity holder of deepPath Inc. and CogniScreen, Inc. He also serves on the scientific advisory board of Altoida Inc. The remaining authors declare no competing interests.

## Acknowledgment

The authors gratefully acknowledge the American Society of Nephrology (ASN) for providing access to the Nephrology Self-Assessment Program (NephSAP) content, which was instrumental in conducting this study. This access enabled a comprehensive evaluation of large language models within the context of nephrology education and clinical knowledge assessment.

## Notes

### Author Declarations

Access to the nephSAP data was granted for the present work after registration to American Society of Nephrology (ASN) on 07/04/2023 with registration ID ASNID#780347. All data were anonymized within main text, figures, and tables.

